# The feasibility of an objective measure of the parent-child relationship in health visiting practice: assessment of the Maternal Postnatal Attachment Scale

**DOI:** 10.1101/2021.11.30.21267061

**Authors:** Abigail Dunn, Philippa K. Bird, Charlotte Endacott, Tracey Bywater, Joanna Howes, Josie Dickerson

## Abstract

Positive parent infant relationships are key to achieving long term child outcomes. Identifying parents who may need support is difficult because of a lack of robust assessment tools. Working in partnership with health services we piloted the Maternal Postnatal Attachment Scale (MPAS) in a deprived, multi-ethnic urban community in Bradford, UK.

The pilot aimed to assess the clinical utility of MPAS to identify need for support: Was it administered to a representative group of women? Is MPAS valid for this population?

Data were linked to a cohort study in the pilot area (Born in Bradford’s Better Start - BiBBS). Chi Square tests assessed sample representativeness (age, ethnicity, parity, English language, education, deprivation). Exploratory factor analysis explored MPAS’ validity.

563 women in BiBBS were eligible, 210 (37%) completed MPAS. No differences were found between completers and non-completers, suggestive of a representative sample. In total, 336 women completed MPAS in the pilot. MPAS had ceiling effects and a satisfactory factor structure could not be identified, indicating poor psychometric properties

Health visitors were successful in administering MPAS to a representative sample, but poor psychometric robustness indicates that MPAS is unsuitable for routine use in this setting. A gap for such a measure remains.

**Statement of relevance to practitioners:** This research shows that:

1. Health practitioners working in universal early years services were willing and able to integrate structured assessments of parent-infant relationship into their routine practice
2. There is no evidence that health professionals were less likely to offer traditionally considered ‘hard to reach’ families an assessment of their parent-infant relationship
3. The Maternal Postnatal Attachment Scale did not demonstrate psychometric robustness when delivered in routine, universal, health visiting services in Bradford as part of an initial assessment of parent-infant relationship, and therefore cannot be recommended for continued routine use.

**Statement of relevance to the field:** This pilot study explores a significant gap in the field, namely how universal services can assess parent-infant relationship to facilitate timely signposting to appropriate services, in a preventative model, to those families that may benefit. This study contributes to the evidence base by assessing if health professionals working in a universal service can offer an assessment to a representative group of families, including families who may be considered hard to reach as well as providing psychometric evidence on the Maternal Postnatal Attachment Scale. We did not find good evidence for the psychometric properties of the Maternal Postnatal Attachment Scale when used in this way in Bradford. We found that health professionals offered the assessment to a representative sample of families, including those who may be at an enhanced risk of health inequalities because of their ethnicity, age, education, and wider socioeconomic circumstances.

This evidence is important for measure/tool selection for community studies. The findings also emphasise that practitioners can integrate tools for assessment in their practice, including with families who are at an increased risk of experiencing inequalities.

**Diversity and anti-racist scholarship:** This study was designed and executed in a very diverse community with approximately 60% of the population identifying as Asian/Asian British: Pakistani, and 10% of the population identifying as White: British and the remaining population identifying with a wide range of ethnicities. The health visiting service deliberately engages with the whole population in a culturally sensitive way, including ensuring staff speak key community languages and using translators as required. We included specific tests of representativeness as part of the study design and found that participants in the study were representative of the wider community in terms of ethnicity and English language comprehension.

## Introduction

### Background

The ability of a mother to interact with her infant sensitively, whilst attuned to their infant’s mental state and level of development is a crucial precursor of a child’s ability to develop a secure attachment (Ainsworth et al., 1978; Kim et al., 2017). Secure attachment predicts a child’s later social-emotional development (Fearon et al., 2010; Le Bas et al., 2020). Insecure attachment prevalence rates are estimated to be high, at 35% in a Danish community study (Skovgaard, 2010), while attachment disorder rates in the UK are 1.4% (Minnis et al., 2013).

The importance of a healthy parent-infant relationship for children’s future development is recognised in the UK in three national clinical guidelines, with assessment, early identification and intervention being key recommendations (NICE guidelines CG192 - antenatal and postnatal mental health (NICE, 2020), NG26 - attachment in children in care (NICE, 2015), and PH40 - social and emotional well-being: the early years (NICE, 2012).

However, despite the clear importance of early identification and intervention NICE guidance acknowledges that no tools have been identified for use for the 0-12 months postnatal period – a critical time point to allow early identification and prevention of issues. The tools recommended by NICE for identification in pre-school children require clinical expertise and observations making them expensive for use in universal services (NG 26, NICE, 2015). Two recent reviews of self-report tools for measuring maternal dimensions of the parent-infant relationship concluded that no available measures could be recommended for use, in the main due to the lack of evidence about the clinical utility and psychometric properties of the tools (Mathews et al., 2019; Wittkowski et al., 2020).

NICE Guidance (NG26) notes this gap in their research recommendations where they state the need to “*Develop reliable and valid screening assessment tools for attachment and sensitivity that can be made available and used in routine health, social care and education settings*”(NICE 2015).

In the UK all children and their parents receive a minimum of 5 mandated visits from a health visitor during pregnancy up to 2.5 years of age to support the child’s safety and development. Early parent-infant relationship is recognised as one of the main priorities for health visiting in Early years high impact area 2: Maternal and family mental health (PHE, 2020). However, as far as we are aware, the majority of health visitors across the UK rely on personal observations and professional judgement to identify issues with the parent-child relationship (Appleton et al., 2013; Wilson et al., 2010). Such assessments are subjective and hard to validate. In addition, such observations are recorded in free text rather than coded sections of a healthcare record making extraction of such assessments on a population level challenging. A lack of validated and coded recording may impact on the chances of high quality, joined up clinical care for mother and baby, and limits the ability of researchers and health organisations to characterise prevalence and epidemiology more accurately; identify local levels of need and plan for service provision.

In Bradford, as a part of the Better Start Bradford programme, (see Box 2), the decision was made to pilot the implementation of an objective and validated assessment tool into universal health visiting practice within an inner-city area of Bradford. Given the lack of recommendations for a tool, the research team worked together with the health visiting service to complete a brief review of potential measures focussing on evidence of validity and reliability as well as potential clinical utility. This review used the same methodology as a larger review by the team (Blower et al, 2019). A number of measures were considered (Brockington et al., 2001; Taylor et al., 2005; Muller, 1994; Hovik et al., 2013; Cuijilts et al., 2016). The Maternal Postnatal Attachment Scale (MPAS) (Condon & Corkindale, 1998) was selected as the best option (from the few existing appropriate measures) for the pilot based on previous research with this measure, its psychometric properties, is freely and easily available.

### The MPAS

The MPAS was developed by John Condon and colleagues in Australia (Condon & Corkindale, 1998). It is a 19-item measure suitable for use with mothers in the first postnatal year. The items are a mixture of forward and reverse scored items with either 2, 3, 4 or 5 answer categories. Each item is equally weighted so some of the item response categories has decimal scoring. The maximum score is 95, and the theoretical minimum is 19. Lower scores indicate more problematic responses. The MPAS does not have validated cut off points for problematic or concerning relationships and is not intended for use as a diagnostic tool on its own, but as a supportive indication within a holistic assessment.

The MPAS has been assessed for validity and has been described as suitable for use in research and clinical practice (Condon & Corkindale, 1998). A sample of 238 women recruited antenatally completed MPAS at three different timepoints (4 weeks, 4 months, and 8 months). Stability of the measure over time was acceptable (all Pearson correlation coefficients significant at p<0.001) and internal consistency of the measure was acceptably high (alphas>0.7). Factor analysis found that the items loaded onto three factors: Quality of attachment, Absence of hostility and Pleasure in interaction (Condon and Corkindale, 1998).

MPAS has not been widely validated, with only five studies which validate the measure (Condon & Corkindale, 1998; Feldstein et al., 2004; Scopesi et al., 2004; van Bussell et al., 2010; Reira-Martin, 2018). These studies were all included in the review by Wittkowski and colleagues (2020) which concluded that the MPAS (and the other included measures) lack evidence of validation, and that if using the measures consideration needs to be given to the robustness of the findings.

The aim of this paper was to assess the clinical utility of the MPAS in universal health visiting services in a disadvantaged and ethnically diverse population.

### Specific objectives were to

- Explore how feasible and acceptable implementation of this tool was within standard health visiting practice in a disadvantaged and ethnically diverse population
- Evaluate the validity and reliability of the tool when used within standard health visiting practice in a disadvantaged and ethnically diverse population

## Methods

### Implementation of the MPAS pilot

The MPAS was piloted as a universal assessment at the 3-4-month health visiting contact, over a 1-year period between 8^th^ May 2017 and 8^th^ May 2018. The 3-4 month contact is not one of the nationally mandated contacts but is an additional universal contact offered in Bradford. For women who did not speak English, there were options of an Urdu translated MPAS, administered by a bilingual health visitor, or support from a bilingual health visitor or interpreter for other languages. Several health visitors in the Better Start Bradford (BSB) area speak community languages and work predominately with families in these languages.

Training on MPAS administration and scoring, and what to do if concerns were identified, was provided by local perinatal mental health specialists. A referral pathway into local perinatal mental health services, discrete interventions and children’s services was developed.

Health visitors were asked to record: if the MPAS was offered; if declined, the reasons for this; whether it was self-completed, completed with the help of the health visitor or with an interpreter; and the language used to complete the tool.

### MPAS sample eligibility

All women with babies, living in the pilot BSB areas, who had a 3–4-month health visitor contact were eligible to complete the MPAS. Of those who were eligible, women who had a reference to the MPAS assessment in their health record were defined as having been offered the assessment. Those who had no record of any questions being completed were defined as not participating, and reasons for non-participation were reviewed.

To assess clinical utility, those who had one or more questions completed in their health record were defined as having participated in the MPAS assessment, and those who completed 15 or more of the 19 questions were defined as having completed the MPAS.

### Pilot Study Eligibility

#### a. Clinical Utility

All women seen by health visitors for a 3-4 month visit within the time period of the pilot (8^th^ May 2017 and 8^th^ May 2018) for whom routine health data was available were included in the analysis of coverage and completion. For the representativeness analysis, BiBBS participants who had an infant aged 3-4 months between the 8^th^ May 2017 and 8^th^ May 2018 (the time period of the pilot study) and were living in the Better Start Bradford area were included.

#### b. Validity & Reliability

The same routine health data used for the coverage and completion analyses were used for the factor analysis. However, participants who did not complete the MPAS in English were excluded, as were all participants who did not complete all 19 questions.

### Data sources

Routine health visitor data for all eligible women was anonymised and shared with the research team.

For the representativeness analysis, data on the characteristics of eligible women in the pilot area were obtained using the Born in Bradford’s Better Start (BiBBS) research cohort (see Box 1). Data included sociodemographic characteristics for all women with infants aged 3-4 months. As part of the BiBBS cohort, routine health visiting data (including MPAS data) were linked to cohort data. This enabled a comparison of women in the cohort who did and did not participate in the MPAS assessment.

### Analysis

#### Objective 1: Feasibility and acceptability of implementation

An acceptable measure would be one which health visitors are willing to ask, and one which women are willing to complete (de Vet et al., 2011). Three key factors were explored, as recommended by de Vet et al (2011):

a. Coverage: What percentage of eligible women took part in the MPAS pilot?
b. Completion: What percentage of eligible women completed the tool? Adequate completion was defined as 85% women or higher completing at least 15/19 questions.
c. Representativeness: Are there differences in the characteristics of women who took part in the pilot and completed the MPAS, compared to women who were eligible but did not take part?

For a) and b), descriptive statistics were calculated for all eligible women (women with a 3–4-month health visitor check during the pilot period) using routine service data. For c), we compared age, ethnicity, English language ability, education and material deprivation in the BiBBS data using a Chi Square test for differences in proportion. Missing data led to a casewise deletion.

#### Objective 2: Validity and reliability of the tool

The content validity of the MPAS was established in the original development study (Condon & Corkindale, 1998). but there is limited evidence about other measurement properties. Therefore, the structural validity and internal consistency of the MPAS were assessed in the pilot using exploratory factor analysis (de Vet et al, 2011; Prinsen et al, 2018; Mokkink et al, 2017; Terwee et al, 2018).

This method used a staged approach that: 1) determined the missingness and variation of scores on individual MPAS items; 2) identified the level of correlation between items, and; 3) provided an interpretation of the structural validity and internal consistency.

In stage 1, items which did not show any variation were identified and removed from the analysis. The remaining items were taken forward into stage 2 where a correlation matrix using Pearson correlation coefficients was constructed. For Structural Validity, any items that did not correlate with at least one other item with a coefficient > 0.2 were identified and removed from the analysis, and similarly any items with a coefficient of > 0.9 were identified and removed. No restrictions were made as to the number of factors to be returned by the analysis.

Assessment of the adequacy of the sample for factor analysis was made using the KMO measure of sampling adequacy which should be above 0.5 and Bartlett’s test of sphericity which should be significant. Exploratory factor analysis was selected because using MPAS in routine service and using MPAS in the UK were both new uses of the tool. Items were required to load onto their factor with a loading of at least 0.5, and items which did not meet this threshold were deleted from the measure item by item. Items which have loading of over 0.3 onto multiple factors in the final measure will be considered for deletion but may be retained Eigenvalues were calculated, and a Scree plot was created to determine the number of factors to retain in the analysis. A threshold of a minimum combined proportion of variance of >50% explained by the factors was set.

For internal consistency Cronbach’s alpha were calculated for each subscale of the above factor analysis, with an expected minimum of 0.7 and of >0.9 being desirable. If the Cronbach’s alpha was not satisfactory, items with poor correlation will be deleted and the Cronbach Alpha recalculated.

All Data were analysed using SPSS (v24).

### Ethics

The MPAS pilot is a service evaluation (HTA decision ref: 60/88/81 February 2017) and therefore no NHS or departmental ethical approval was required. The BiBBS study received ethical approval by Bradford Leeds NHS Research Ethics Committee (15/YH/0455), and research governance approval from Bradford Teaching Hospitals NHS Foundation Trust. All data were anonymised prior to analysis and are stored securely at the Bradford NHS Teaching Hospital.

## Results

### Objective 1: Feasibility and acceptability of implementation

During the study period, 37 health visitors were working in the pilot area and 833 women had a 3–4-month visit. In total, 35 of 37 (95%) health visitors completed at least one MPAS assessment, and the number completed per health visitor ranged from 1 to 66.

Of the 833 eligible women, 435 (52%) had been offered the MPAS and of these, 347 (42% of total eligible women and 80% of those offered the MPAS) women participated in the assessment. Reasons for not participating included refusal, having a conversation instead of using the measure, another person present, and inadequate time. Of the 347 who participated, 302 (87%) completed the assessment.

563 BiBBS participants were eligible for this study. Of these women, 210 had been offered the MPAS assessment. There were no significant differences between women who were and were not offered an MPAS for any of the characteristics examined (See Table1).

**Table 1:**
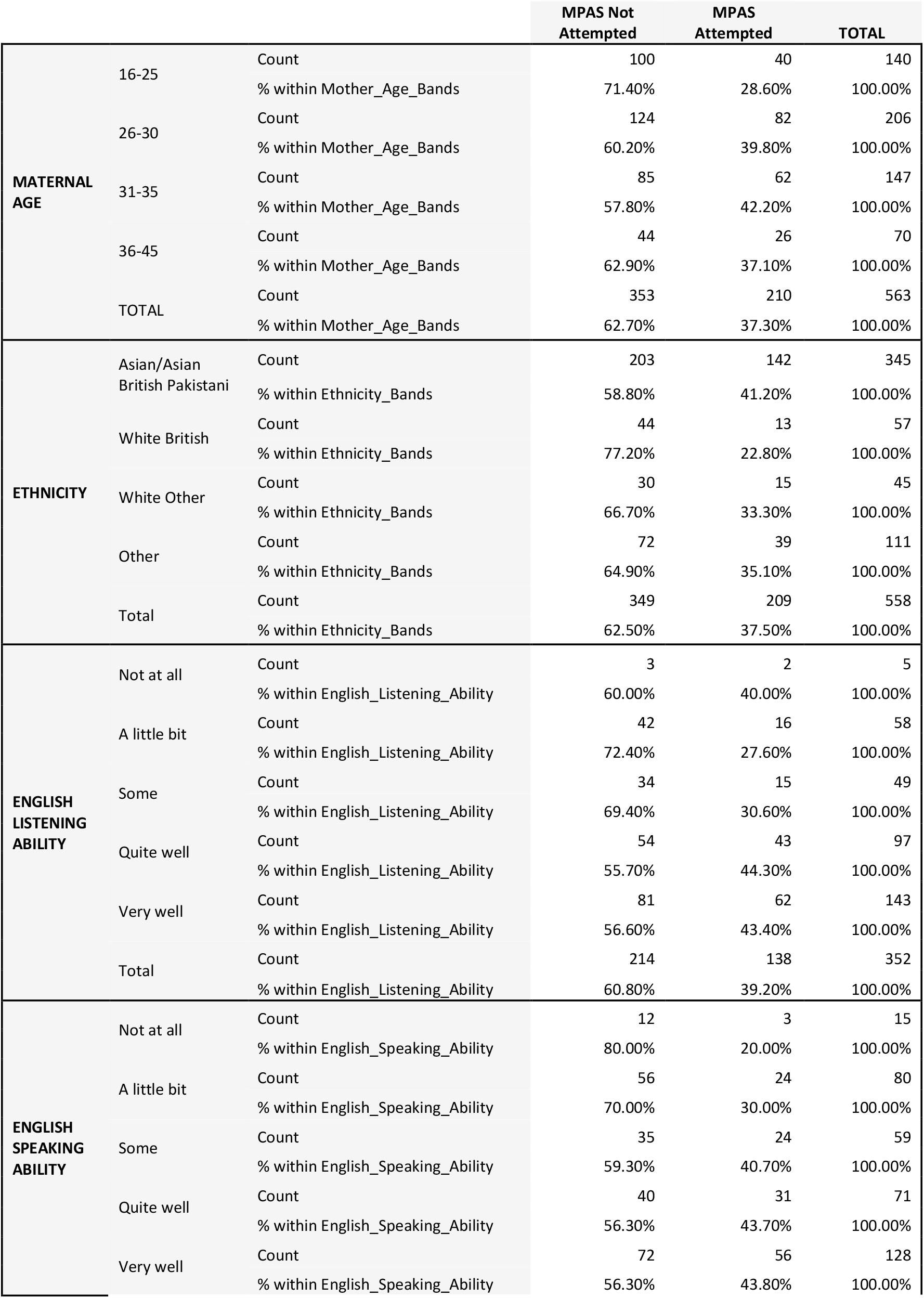
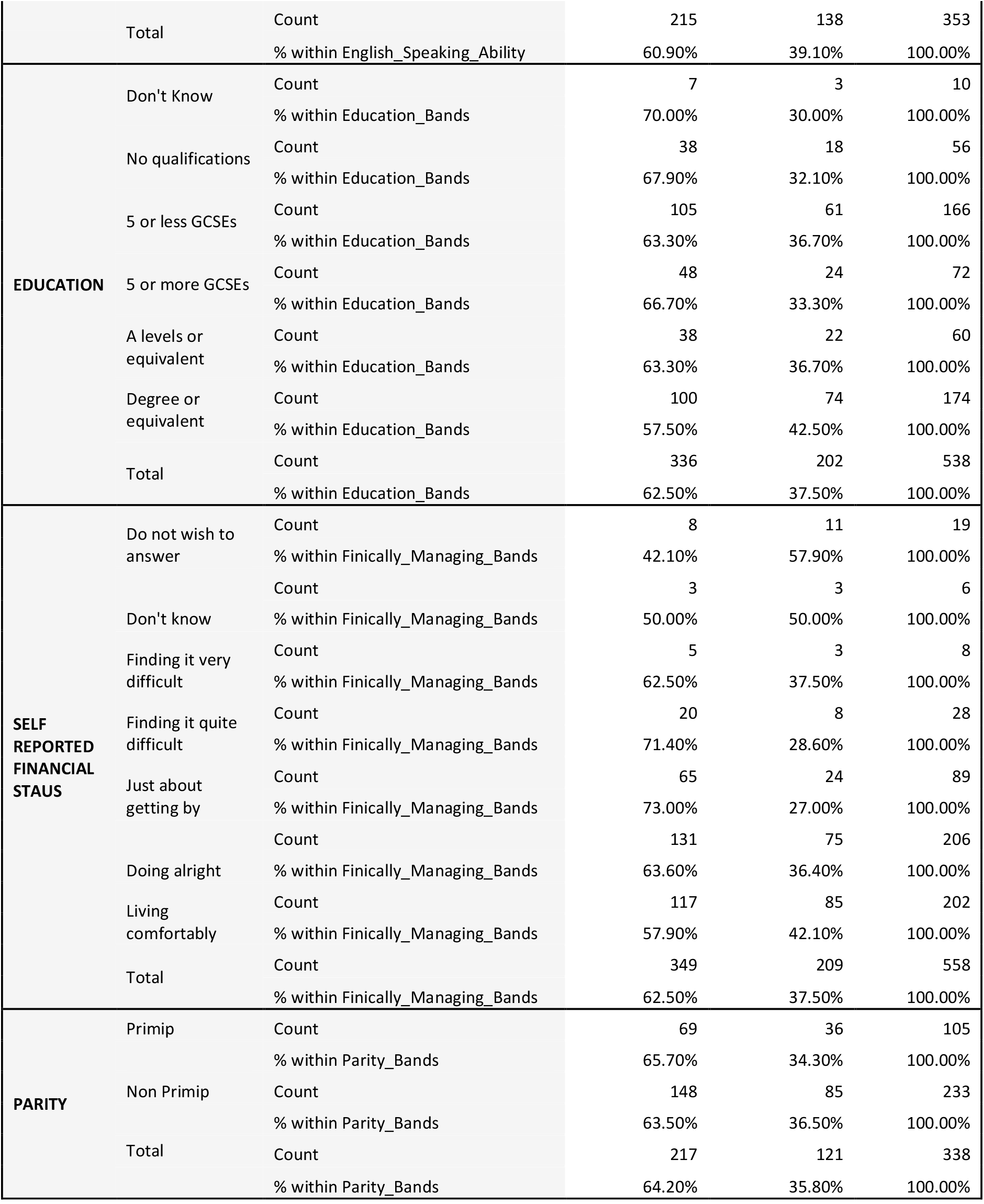
MPAS attempted with socio-demographics.

### Objective 2: Validity and reliability of the tool

198 MPAS assessments were available for the factor analysis (see Figure1). Overall, MPAS scores were skewed, with the vast proportion of women scoring very high, indicative of no concern (Figure 2). 21% of women who completed the MPAS in English scored the maximum score of 95 on the tool.

**Figure 1:**
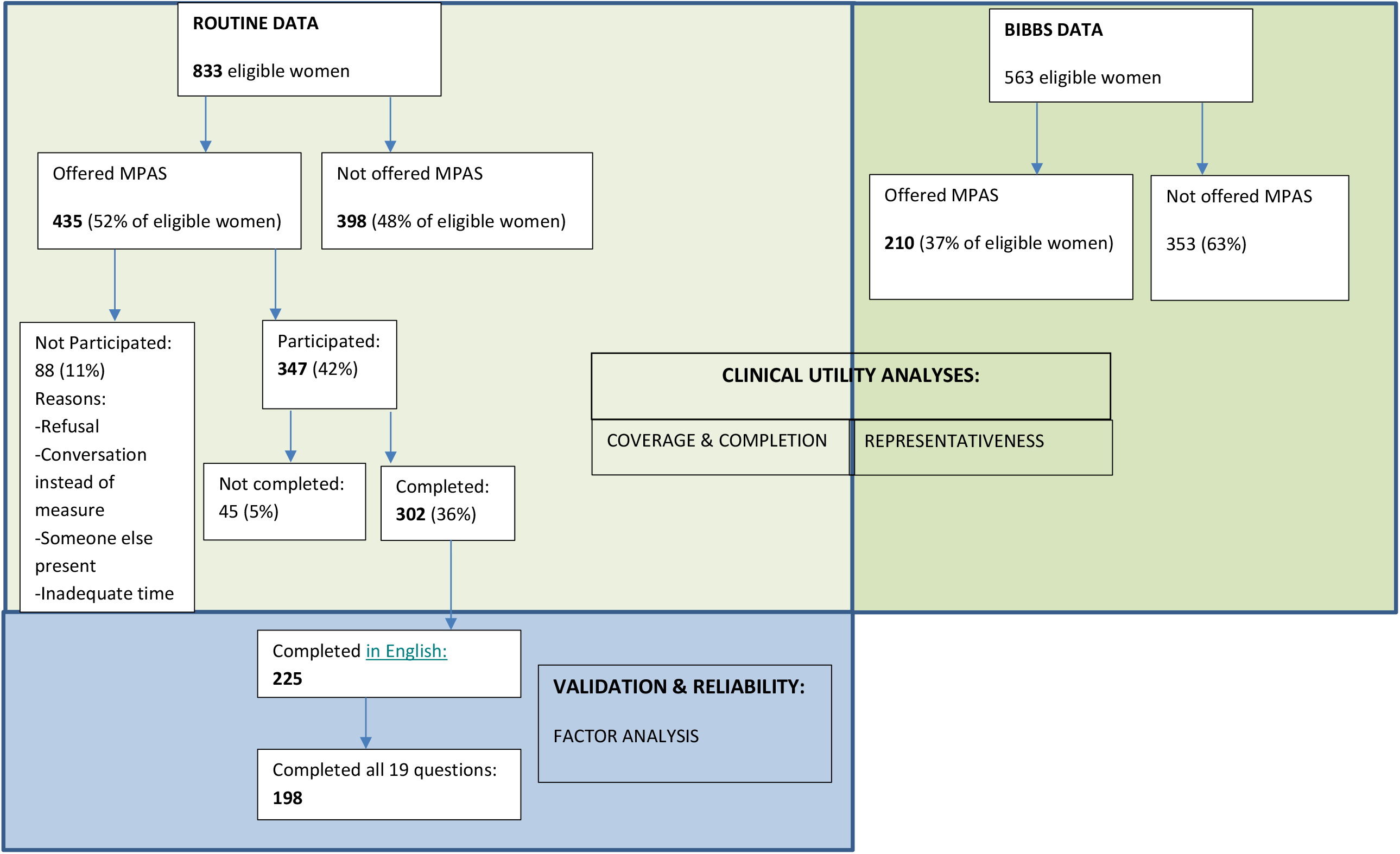
Flow of participants in the MPAS pilot study broken down by the samples used in the clinical utility and validation analyses.

**Figure 2:**
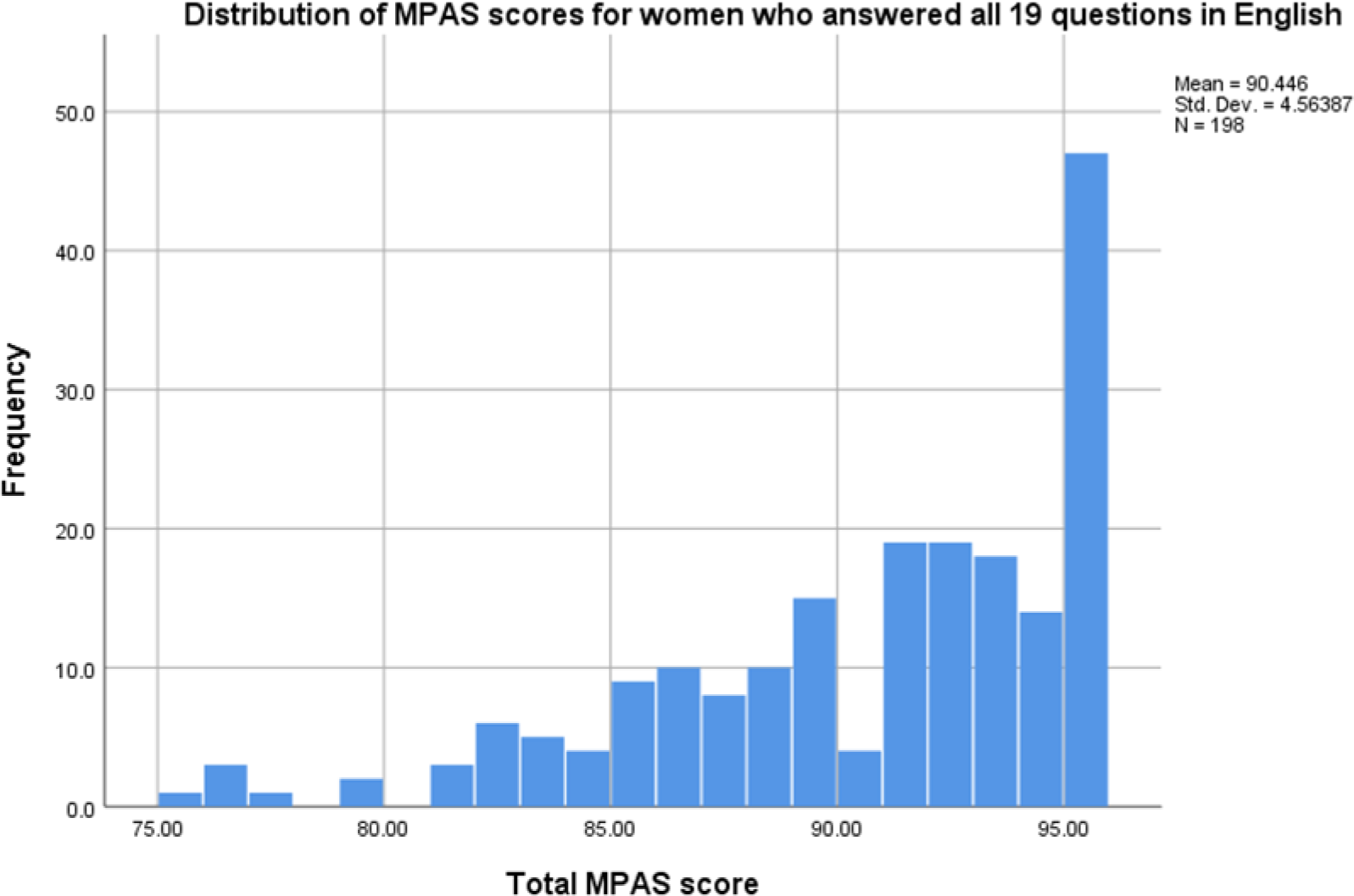
Distribution of MPAS scores for women who answered all 19 questions in English.

Item response was high. The highest proportion of observed missing data (in 6.7% of cases) was for question 9 (“When I leave the baby…”). This item level missingness is not high enough to suggest that the item should be dropped (de Vet et al, 2011).

Item variation identified that for 9 of the 19 questions, at least one of the response categories was not used by any of the participants. In the case of question 14 “I now think of the baby as…”, all 198 women selected the response “very much my own baby”. Due to the lack of variation in response in question 14 this item was dropped from the next stage of analysis.

Table 2 shows the correlation matrix of the remaining 18 items of the MPAS. Items 7 (“When I am with the baby and other people are present, I feel proud of the baby…”) and 12 (“When I am with the baby: … try to prolong time I spend with…”) were not correlated with any of the other MPAS items with correlation coefficients <0.2 and were removed from further analysis. The factor analysis moved forward with 16 items (i.e. without question 7,12 and 14).

**Table 2:** Correlation matrix of 18 MPAS items.

| Question number | 1 | 2 | 3 | 4 | 5 | 6 | 7 | 8 | 9 | 10 | 11 | 12 | 13 | 15 | 16 | 17 | 18 | 19 |
| --- | --- | --- | --- | --- | --- | --- | --- | --- | --- | --- | --- | --- | --- | --- | --- | --- | --- | --- |
| 1 | 1 | 0.161 | 0.056 | 0.207 | 0.244 | 0.211 | 0.048 | -0.059 | 0.283 | 0.166 | 0.108 | 0.055 | 0.072 | 0.127 | 0.287 | 0.273 | 0.134 | 0.177 |
| 2 | 0.161 | 1 | 0.208 | 0.123 | 0.056 | 0.137 | 0.102 | -0.028 | 0.128 | 0.085 | 0.137 | 0.097 | 0.016 | 0.066 | 0.078 | -0.020 | -0.009 | 0.016 |
| 3 | 0.056 | 0.208 | 1 | 0.249 | -0.034 | -0.028 | -0.083 | -0.034 | 0.170 | 0.111 | 0.242 | 0.086 | 0.444 | 0.186 | 0.065 | 0.060 | 0.251 | 0.111 |
| 4 | 0.207 | 0.123 | 0.249 | 1 | 0.156 | 0.164 | -0.071 | -0.042 | 0.026 | 0.152 | 0.053 | -0.034 | -0.015 | 0.102 | 0.177 | 0.280 | 0.018 | 0.363 |
| 5 | 0.244 | 0.056 | -0.034 | 0.156 | 1 | 0.320 | 0.014 | -0.045 | 0.236 | 0.199 | 0.218 | 0.110 | 0.027 | 0.111 | 0.193 | 0.218 | 0.216 | 0.175 |
| 6 | 0.211 | 0.137 | -0.028 | 0.164 | 0.320 | 1 | -0.004 | -0.038 | -0.018 | 0.070 | 0.019 | -0.031 | 0.024 | 0.067 | 0.063 | 0.061 | 0.124 | 0.086 |
| 7 | 0.048 | 0.102 | -0.083 | -0.071 | 0.014 | -0.004 | 1 | -0.045 | 0.112 | 0.078 | -0.035 | -0.037 | 0.131 | 0.168 | 0.088 | 0.105 | -0.074 | 0.061 |
| 8 | -0.059 | -0.028 | -0.034 | -0.042 | -0.045 | -0.038 | -0.045 | 1 | 0.031 | -0.033 | -0.053 | -0.013 | -0.034 | 0.214 | 0.024 | -0.050 | 0.031 | -0.041 |
| 9 | 0.283 | 0.128 | 0.170 | 0.026 | 0.236 | -0.018 | 0.112 | 0.031 | 1 | 0.188 | 0.361 | 0.075 | 0.189 | 0.104 | 0.131 | 0.066 | 0.237 | 0.145 |
| 10 | 0.166 | 0.085 | 0.111 | 0.152 | 0.199 | 0.070 | 0.078 | -0.033 | 0.188 | 1 | 0.224 | -0.027 | 0.105 | 0.202 | 0.225 | 0.135 | 0.189 | 0.368 |
| 11 | 0.108 | 0.137 | 0.242 | 0.053 | 0.218 | 0.019 | -0.035 | -0.053 | 0.361 | 0.224 | 1 | 0.184 | 0.295 | 0.082 | 0.184 | -0.027 | 0.181 | 0.221 |
| 12 | 0.055 | 0.097 | 0.086 | -0.034 | 0.110 | -0.031 | -0.037 | -0.013 | 0.075 | -0.027 | 0.184 | 1 | 0.082 | -0.038 | 0.198 | -0.041 | 0.182 | -0.033 |
| 13 | 0.072 | 0.016 | 0.444 | -0.015 | 0.027 | 0.024 | 0.131 | -0.034 | 0.189 | 0.105 | 0.295 | 0.082 | 1 | 0.037 | 0.085 | 0.068 | 0.237 | 0.076 |
| 15 | 0.127 | 0.066 | 0.186 | 0.102 | 0.111 | 0.067 | 0.168 | 0.214 | 0.104 | 0.202 | 0.082 | -0.038 | 0.037 | 1 | 0.324 | 0.295 | 0.052 | 0.202 |
| 16 | 0.287 | 0.078 | 0.065 | 0.177 | 0.193 | 0.063 | 0.088 | 0.024 | 0.131 | 0.225 | 0.184 | 0.198 | 0.085 | 0.324 | 1 | 0.239 | 0.275 | 0.109 |
| 17 | 0.273 | -0.020 | 0.060 | 0.280 | 0.218 | 0.061 | 0.105 | -0.050 | 0.066 | 0.135 | -0.027 | -0.041 | 0.068 | 0.295 | 0.239 | 1 | 0.166 | 0.194 |
| 18 | 0.134 | -0.009 | 0.251 | 0.018 | 0.216 | 0.124 | -0.074 | 0.031 | 0.237 | 0.189 | 0.181 | 0.182 | 0.237 | 0.052 | 0.275 | 0.166 | 1 | 0.121 |
| 19 | 0.177 | 0.016 | 0.111 | 0.363 | 0.175 | 0.086 | 0.061 | -0.041 | 0.145 | 0.368 | 0.221 | -0.033 | 0.076 | 0.202 | 0.109 | 0.194 | 0.121 | 1 |

\* Correlations between items of higher than 0.2 are highlighted

Examination of the scree plot (Figure 3) shows that there is an indication that a three-factor solution similar to that identified by Condon et al (1998), may be relevant in this population, however, this three-factor solution only explained 41% of the variance. A six factor solution based on all factors with an eigenvalue of >1 explains 67% of the variance. However, on further examination, (table 3x), factor six has only one variable loading onto it. Removing this variable (question 2), another variable (question 9) no longer loads onto the factor solution, leaving factor five with one variable loading onto it (question 4). Removing these three variables leads to a 10-item scale with a four factor solution that explains 60% of the variance. Whilst this solution meets the KMO test for sampling adequacy, and Bartlett’s test of sphericity, interpreting the factor structure highlights that, as well as a relatively large number of factors from just ten items, there are three items (8,15,18) which are loading (at >0.3, but less than <0.5) onto multiple factors impairing interpretation of the factor structure of the tool. As no meaningful factors can be extracted from the MPAS data there is no ability to assess the internal consistency of the extracted factors.

**Table 3:** Factor loadings in six factor solution.

| Component | Total | Initial Eigenvalues |  |
| --- | --- | --- | --- |
|  |  | % of Variance | Cumulative % |
| 1 | 2.692 | 20.706 | 20.706 |
| 2 | 1.445 | 11.117 | 31.823 |
| 3 | 1.254 | 9.647 | 41.471 |
| 4 | 1.179 | 9.069 | 50.539 |
| 5 | 1.093 | 8.408 | 58.947 |
| 6 | 1.042 | 8.016 | 66.963 |
| 7 | .849 | 6.530 | 73.492 |
| 8 | .750 | 5.773 | 79.265 |
| 9 | .684 | 5.259 | 84.524 |
| 10 | .583 | 4.483 | 89.007 |
| 11 | .575 | 4.424 | 93.431 |
| 12 | .501 | 3.851 | 97.282 |
| 13 | .353 | 2.718 | 100.000 |

**Figure 3:**
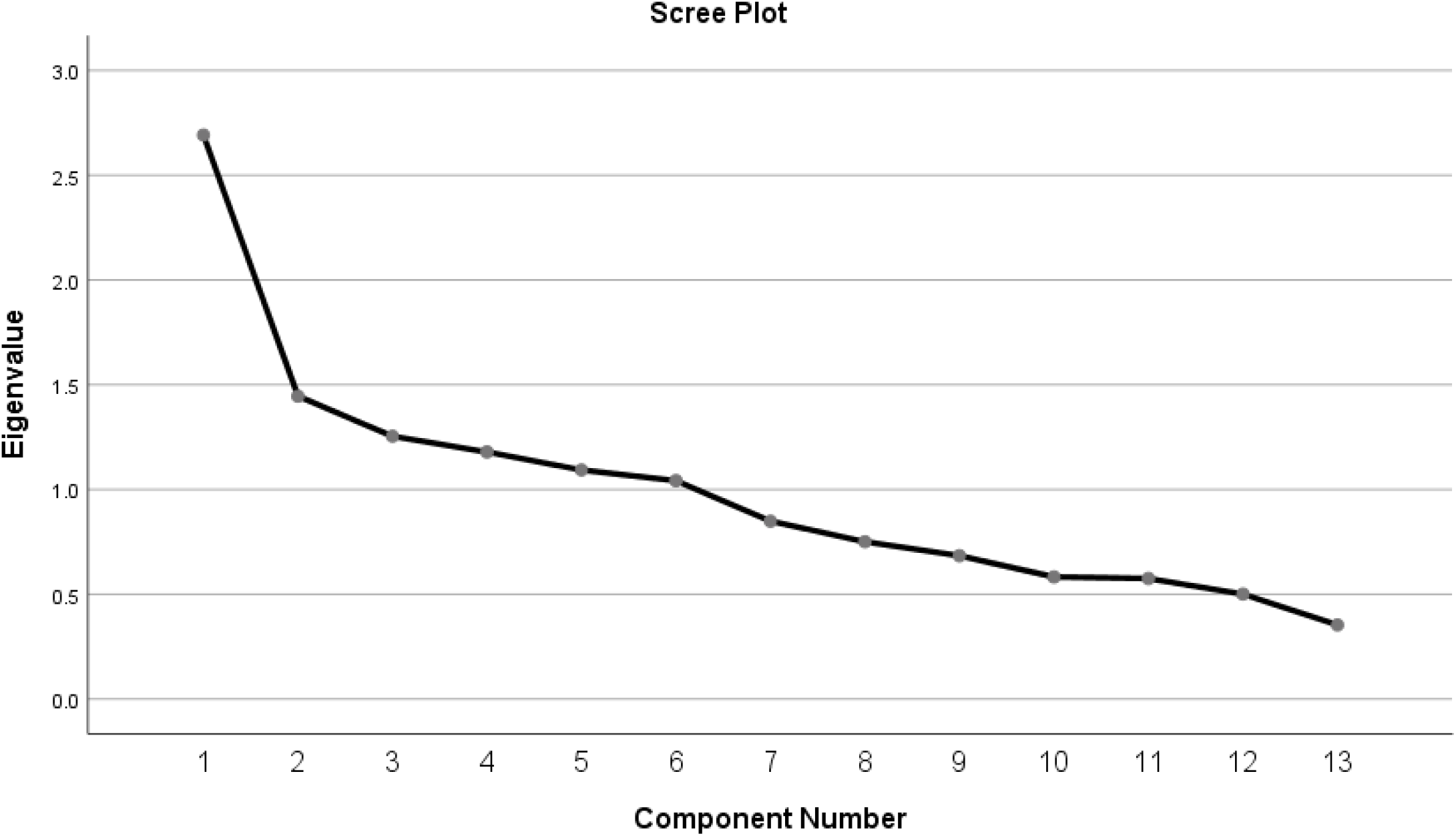
Scree plot.

## Discussion

This study assessed the clinical utility, validity and reliability of the MPAS in universal health visiting services in Bradford. This is the first time that this assessment tool has been used in clinical practice anywhere (to the authors’ knowledge), and in a disadvantaged and ethnically diverse population. In the pilot, health visitors’ use of the tool was inconsistent and only 52% of eligible women were offered an MPAS assessment by the health visitor at the 3-4-month visit. There was considerable variation between health visitors in how often they used the MPAS with their case load, with some health visitors never recording using it, and one using the tool 66 times during the pilot. There were no socio-demographic differences in who was and was not offered the MPAS, suggesting that health visitors were not being biased in who they offered the assessment to. Of those offered the MPAS, 80% participated and 87% completed suggesting that, when offered, it is acceptable. However, the distribution of the scores was highly skewed with little variance and no indication of any concerns detected in the scores. Furthermore, the analysis on the validity of the MPAS tool in this population failed to find evidence of internal consistency or structural validity. The findings suggest that women in the pilot study population did not interpret or respond to the MPAS questions as intended.

Further in-depth exploration of these findings is required to understand why some health visitors used the tool inconsistently, and what the barriers were to completing the tool with almost half of the eligible population. It is important to understand whether the barriers related to the design of the tool or to contextual factors that could be addressed to improve uptake.

Further exploration is also required to understand the lack of variance in the scores in the pilot study. Whilst this could relate to a lack of validity with women perhaps not understanding or interpreting the questions as intended due to cultural and/or language differences. There may however also be reluctance for women to disclose concerns about their relationship with their baby to health professionals. Previous research completed with a similar population has shown that women from ethnic minorities are less likely to have their perinatal mental health identified by health professionals due to a complex interplay of reluctance to disclose (e.g. due to stigma, fear of having their baby taken away), difficulty in identification by health professionals (e.g. use of interpreters, lack of time etc.,) and problems in capturing issues on IT systems (Prady et al, 2021, Prady et al, 2016a., Prady et al, 2016b).

An additional research study has explored these explanations using qualitative interviews with health visitors during this pilot study. The linked paper by Bird et al explores these issues further. Key findings from this paper suggest that although health visitors welcomed the opportunity to discuss the parent infant relationship and there were benefits to using a structured tool, there were also considerable challenges that hindered implementation of the MPAS in a valid and reliable way. Health visitors had concerns around the length of time required to administer the tool, the complexity of the language and the intrusiveness of some questions. These concerns were exacerbated when translation was used. The context that health visitors are working in and lack of time for home visits also posted challenges. Together, the papers highlight the need for a robust, valid measure to assess parent-child relationships in routine practice, with coproduction to ensure clinical utility and acceptability.

Strengths of this study include the evaluation of the use of the MPAS in routine health visiting practice, meaning that findings relate to ‘real world’ use of the measure. The evaluation builds on a successful partnership between the service and evaluation teams, from working together to identify a suitable measure through to evaluation and implementation of findings into practice. This meant that the evaluation considered both theoretical and operational perspectives.

There are two key limitations to this study. Firstly, the BSB population has an unusual profile. The population in BSB are very ethnically diverse (only 10% of the women giving birth in the area identify as White British) and economically deprived, live in an urban area, and are not representative of the wider UK population. As such the findings may not be valid for less deprived, less ethnically diverse, or more suburban/rural communities. It is vital that any objective tool is feasible to implement and meaningful to use with all women and health professionals.

Secondly, the routine setting of the study meant we were reliant on health visiting data, not all of which we had full access to. We had no information about the 50% of eligible women who had contact with a health visitor but who had no information recorded as to if they were asked to complete MPAS. Not knowing why MPAS was not asked in these cases limits our ability to understand how acceptable and useful the MPAS was to both women and health visitors. These limitations mean that caution must be exercised when generalising the findings of the BSB pilot.

## Conclusions

The MPAS was administered to a representative sample by health visitors, but acceptability was low, and the MPAS had poor psychometric properties. Qualitative research (Bird et al. submitted) confirms that the MPAS was not fully understood by the sample, rendering it unacceptable for the Bradford context. Although health visitors welcomed the opportunity to discuss the parent infant relationship, there were also considerable challenges. This included concerns around the complexity and length of the tool itself and the time-pressured context that health visitors are working in.

## Implications for practice and/or further research

Based on the findings from this paper, and Bird et al, (submitted), the gap for a robust, valid measure to assess parent-child relationships in routine practice remains, at least in Bradford. Considering this, we have coproduced a tool with health visitors, service staff and with input from parents, based on the learning from this pilot, and are testing it in routine care.

## Data Availability

Data is stored securely by Born in Bradford at the Bradford Institute for Health Research (BIHR). Data sharing is not applicable to this article. However, please note that data and samples collected throughout the course of the BiBBS cohort will be available to external researchers and proposals for collaboration will be welcomed. Information on how to access the data can be found at: www.borninbradford.nhs.uk.

## Acknowledgements

This study has received funding through a peer review process from the National Lottery Community Fund as part of the A Better Start programme. The funders have not had any involvement in the design or writing of the paper. Authors PKB, AD, JD and TB were supported by the NIHR CLAHRC Yorkshire and Humber (www.clahrc-yh.nihr.ac.uk). JD and TB were supported by the NIHR ARC Yorkshire and Humber (https://www.arc-yh.nihr.ac.uk/). The authors acknowledge the help of Kathryn Willan and Joyti Panesar-Sharma in the preparation of the dataset used. The views and opinions expressed are those of the author(s), and not necessarily those of the NHS, the NIHR or the Department of Health and Social Care.

## Notes

**Conflict of interest statement:** The authors report no conflict of interest

### Competing Interest Statement

The authors have declared no competing interest.

### Author Declarations

The Health Research Authority confirmed that the pilot of the Maternal Postnatal Attachment Scale is considered to be service evaluation, not research, and as such does not require review by an NHS Research Ethics Committee (HRA decision 60/88/81). The Born in Bradfords Better Start study used for some comparative analysis received ethical approval by Bradford Leeds NHS Research Ethics Committee (15/YH/0455), and research governance approval from Bradford Teaching Hospitals NHS Foundation Trust. All data were anonymised prior to analysis and are stored securely at the Bradford NHS Teaching Hospital.

## References

Ainsworth, M., Blehar, M., Waters, E., & Wall, S. N. (1978). Patterns of attachment : a psychological study of the strange situation. Psychology Press.

Appleton, J. V., Harris, M., Oates, J., & Kelly, C. (2013). Evaluating health visitor assessments of mother-infant interactions: a mixed methods study. International Journal of Nursing Studies, 50(1), 5–15. https://doi.org/10.1016/j.ijnurstu.2012.08.008

Bird, P. K., Hindson, Z., Dunn, A., de Chavez, A. C., Dickerson, J., Howes, J & Bywater, T. (2021). Implementing the Maternal Postnatal Attachment Scale in universal services: Qualitative interviews with health visitors. (Submitted)

Blower, S., Gridley, N., Dunn, A., Bywater, T., Hindson, Z. & Bryant, M. (2019). Psychometric Properties of Parent Outcome Measures Used in RCTs of Antenatal and Early Years Parent Programs: A Systematic Review. (2019). Clinical Child and Family Psychology Review. 22, 367–387. https://doi.org/10.1007/s10567-019-00276-2.

Brockington, I. F., Oates, J., George, S., Turner, D., Vostanis, P., Sullivan, M., … Murdoch, C. (2001). A Screening Questionnaire for mother-infant bonding disorders. Archives of Women’s Mental Health, 3(4), 133–140. https://doi.org/10.1007/s007370170010

Condon, J. T., & Corkindale, C. J. (1998). The assessment of parent-to-infant attachment: Development of a self-report questionnaire instrument. Journal of Reproductive and Infant Psychology, 16(1), 57–76. https://doi.org/10.1080/02646839808404558

Cuijlits, I., Wetering AP, V. D., Potharst, E., Truijens, S., van Baar, A., & Vjm, P. (2016). Development of a pre- and postnatal bonding scale (PPBS). Journal of Psychology & Psychotherapy, 6(5).

De Vet, H. C., Terwee, C. B., Mokkink, L. B., & Knol, D. L. (2011). Measurement in medicine: a practical guide. Cambridge university press.

Dickerson, J., Bird, P. K., McEachan, R. R., Pickett, K. E., Waiblinger, D., Uphoff, E., … Wright, J. (2016). Born in Bradford’s Better Start: an experimental birth cohort study to evaluate the impact of early life interventions. BMC Public Health, 16(1), 711. https://doi.org/10.1186/s12889-016-3318-0

Dickerson, J., Bridges, S., Willan, K. et al., Interim Cohort Profile: Born in Bradford’s Better Start (BiBBS) Interventional Birth Cohort Study, the Pre-Covid-19 sample. In prep.

Fearon, R. P., Bakermans-Kranenburg, M. J., van Ijzendoorn, M. H., Lapsley, A. M., & Roisman, G. I. (2010). The significance of insecure attachment and disorganization in the development of children’s externalizing behavior: a meta-analytic study. Child Development, 81(2), 435–456. https://doi.org/10.1111/j.1467-8624.2009.01405.x

Feldstein, S., Hane, A. A., Morrison, B. M., & Huang, K. Y. (2004). Relation of the Postnatal Attachment Questionnaire to the Attachment Q-Set. Journal of Reproductive and Infant Psychology, 22(2), 111–121. https://doi.org/10.1080/0264683042000205972

Høivik, M. S., Burkeland, N. A., Linaker, O. M., & Berg-Nielsen, T. S. (2013). The Mother and Baby Interaction Scale: a valid broadband instrument for efficient screening of postpartum interaction? A preliminary validation in a Norwegian community sample [https://doi.org/10.1111/j.1471-6712.2012.01060.x]. Scandinavian Journal of Caring Sciences, 27(3), p733-739. https://doi.org/https://doi.org/10.1111/j.1471-6712.2012.01060.x

IBM Corp. Released 2016. IBM SPSS Statistics for Windows, Version 24.0. Armonk, NY: IBM Corp.

Kim, B.-R., Chow, S.-M., Bray, B., & Teti, D. M. (2017). Trajectories of mothers’ emotional availability: relations with infant temperament in predicting attachment security. Attachment & human development, 19(1), 38–57. https://doi.org/10.1080/14616734.2016.1252780

Le Bas, G. A., Youssef, G. J., Macdonald, J. A., Rossen, L., Teague, S. J., Kothe, E. J., … Hutchinson, D. M. (2020). The role of antenatal and postnatal maternal bonding in infant development: A systematic review and meta-analysis [https://doi.org/10.1111/sode.12392]. Social Development, 29(1), 3-20. https://doi.org/ https://doi.org/10.1111/sode.12392

Mathews, T. L., Emerson, M. R., Moore, T. A., Fial, A., & Hanna, K. M. (2019). Systematic Review: Feasibility, Reliability, and Validity of Maternal/Caregiver Attachment and Bonding Screening Tools for Clinical Use. Journal of Pediatric Health Care, 33(6), 663–674. https://doi.org/https://doi.org/10.1016/j.pedhc.2019.04.018

Minnis, H., Macmillan, S., Pritchett, R., Young, D., Wallace, B., Butcher, J., … Gillberg, C. (2013). Prevalence of reactive attachment disorder in a deprived population. British Journal of Psychiatry, 202(5), 342–346. https://doi.org/doi:10.1192/bjp.bp.112.114074

Mokkink, L. B., de Vet, H. C. W., Prinsen, C. A. C., Patrick, D. L., Alonso, J., Bouter, L. M., & Terwee, C. B. (2018). COSMIN Risk of Bias checklist for systematic reviews of Patient-Reported Outcome Measures. Quality of life research : an international journal of quality of life aspects of treatment, care and rehabilitation, 27(5), 1171–1179. https://doi.org/10.1007/s11136-017-1765-4

Müller, M. E. A Questionnaire to Measure Mother-to-Infant Attachment. (1994). J Nurs Meas(2), 129–141. https://doi.org/10.1891/1061-3749.2.2.129

NICE. (2012). Public Health Guidance PH40. Social and emotional wellbeing: early years. https://www.nice.org.uk/guidance/ph40

NICE. (2015). Children’s attachment: attachment in children and young people who are adopted from care, in care or at high risk of going into care. Retrieved 27 Nov 2017 from https://www.nice.org.uk/guidance/ng26

NICE. (2020). Clinical guideline CG192. Antenatal and postnatal mental health: clinical management and service Guidance. https://www.nice.org.uk/guidance/cg192.

Prady, S., Endacott, C., Dickerson, J., Bywater, T. J., & Blower, S. L. (2021). Inequalities in the identification and management of common mental disorders in the perinatal period: An equity focused re-analysis of a systematic review. PLoS Online. 16(3), e0248631. https://doi.org/10.1371/journal.pone.0248631

Prady, S.L., Pickett, K.E., Petherick, E.S., Gilbody, S., Croudace, T., Mason, D., Sheldon, T.A. & Wright, J. (2016a). Evaluation of ethnic disparities in detection of depression and anxiety in primary care during the maternal period: combined analysis of routine and cohort data. Br J Psychiatry. 208(5):453–61. https://doi.org/10.1192/bjp.bp.114.158832 PMID: 26795424

Prady, S.L., Pickett, K.E., Gilbody, S., Petherick, E.S., Mason, D., Sheldon, T.A. & Wright, J. (2016b). Variation and ethnic inequalities in treatment of common mental disorders before, during and after pregnancy: combined analysis of routine and research data in the Born in Bradford cohort. BMC Psychiatry.16:99. https://doi.org/10.1186/s12888-016-0805-x PMID: 27071711

Prinsen, C. A. C., Mokkink, L. B., Bouter, L. M., Alonso, J., Patrick, D. L., de Vet, H. C. W., & Terwee, C. B. (2018). COSMIN guideline for systematic reviews of patient-reported outcome measures. Quality of life research : an international journal of quality of life aspects of treatment, care and rehabilitation, 27(5), 1147–1157. https://doi.org/10.1007/s11136-018-1798-3

Public Health England. (2020). Early years high impact area two: Supporting good parental mental health. https://assets.publishing.service.gov.uk/government/uploads/system/uploads/attachment_data/file/942475/Maternity_high_impact_area_2_Supporting_good_parental_mental_health.pdf

Riera-Martín, A., Oliver-Roig, A., Martínez-Pampliega, A., Cormenzana-Redondo, S., Clement-Carbonell, V., & Richart-Martínez, M. (2018). A single Spanish version of maternal and paternal postnatal attachment scales: validation and conceptual analysis. PeerJ, 6, e5980. https://doi.org/10.7717/peerj.5980

Scopesi, A., Viterbori, P., Sponza, S., & Zucchinetti, P. (2004). Assessing mother-to-infant attachment: the Italian adaptation of a self-report questionnaire. Journal of Reproductive and Infant Psychology, 22(2), 99–109. https://doi.org/10.1080/0264683042000205963

Skovgaard, A. M. (2010). Mental health problems and psychopathology in infancy and early childhood. An epidemiological study. DanMedBull, 57(10).

Taylor, A., Atkins, R., Kumar, R., Adams, D., & Glover, V. (2005). A new Mother-to-Infant Bonding Scale: links with early maternal mood. Arch Womens Ment Health, 8(1), 45–51. https://doi.org/10.1007/s00737-005-0074-z

Terwee, C. B., Prinsen, C. A. C., Chiarotto, A., Westerman, M. J., Patrick, D. L., Alonso, J., … Mokkink, L. B. (2018). COSMIN methodology for evaluating the content validity of patient-reported outcome measures: a Delphi study. Quality of Life Research, 27(5), 1159–1170. https://doi.org/10.1007/s11136-018-1829-0

van Bussel, J. C., Spitz, B., & Demyttenaere, K. (2010). Three self-report questionnaires of the early mother-to-infant bond: reliability and validity of the Dutch version of the MPAS, PBQ and MIBS. Archives of Women’s Mental Health, 13(5), 373–384. https://doi.org/10.1007/s00737-009-0140-z

Wilson, P., Thompson, L., Puckering, C., McConnachie, A., Holden, C., Cassidy, C., Gillberg, C. (2010). Parent-child relationships: are health visitors’ judgements reliable? Community Practitioner, 85(2), 22–25.

Wittkowski, A., Vatter, S., Muhinyi, A., Garrett, C., & Henderson, M. (2020). Measuring bonding or attachment in the parent-infant-relationship: A systematic review of parent-report assessment measures, their psychometric properties and clinical utility. Clinical Psychology Review, 82, 101906. https://doi.org/https://doi.org/10.1016/j.cpr.2020.101906

